# Objective dystonia prediction with MRI after neonatal hypoxic-ischemic encephalopathy

**DOI:** 10.1101/2020.05.29.20116947

**Authors:** Keerthana Chintalapati, Hanyang Miao, Amit Mathur, Jeff Neil, Bhooma R. Aravamuthan

## Abstract

**Aim:** To determine an objective and clinically-feasible method to predict dystonia in cerebral palsy (CP) using magnetic resonance imaging (MRI) following neonatal hypoxic-ischemic encephalopathy (HIE).

**Methods:** In this retrospective case-control study, we examined brain MRIs in neonates at age 4–5 days who underwent therapeutic hypothermia for HIE at a single tertiary care center. The lower average apparent diffusion coefficient (ADC) values between the left and right striatum and thalamus were determined using clinically-integrated software (IBM iConnect Access). Neonatal neurology, movement disorder, and cerebral palsy specialist notes were screened through age 5 years for motor abnormality documentation.

**Results:** In 50 subjects, ADC values significantly predicted dystonia in CP with receiver operator characteristic areas under the curve of 0.862 (p = 0.0004) in the striatum and 0.838 (p = 0.001) in the thalamus. Striatal ADC values less than 1.014×10^−3^ mm^2^/s provided 100% specificity and 70% sensitivity for dystonia. Thalamic ADC values less than 0.973×10^−3^ mm^2^/s provided 100% specificity and 80% sensitivity for dystonia.

**Interpretation:** In this small retrospective study, analysis of clinically-acquired MRIs predicted dystonia with high specificity following neonatal HIE. This could be a useful prognostication adjunct guiding when to establish appropriate vigilance for dystonia in CP.

## Introduction

Assessment of brain injury severity on magnetic resonance imaging (MRI) is often used for motor prognostication following neonatal hypoxic-ischemic encephalopathy (HIE).^1,2^ Validated MRI brain injury scales, while academically valuable, require subjective review by expert consensus^1–4^ and thereby may not be practical for real-time clinical prognosticiation. Furthermore, the injury patterns and severity predisposing to different types of motor impairment, such as dystonia versus spasticity in cerebral palsy (CP), are unknown. Studies examining injury patterns associated with dystonia in CP have been difficult to generalize to clinical practice due to lack of differentiation between different CP etiologies, failure to employ standardized and objective image analysis methods, and lack of specialist determination of motor phenotypes^5^. Therefore, it remains challenging to accurately and objectively use MRI in routine clinical practice for motor prognosis following neonatal brain injury.

We sought to begin developing an objective way to predict dystonia, spasticity, or normal motor development in infants who underwent therapeutic hypothermia for neonatal HIE by analyzing brain MRIs obtained during routine clinical care using only clinically-available image viewing software. Noting that the deep grey matter injury has been associated with dystonia in CP^6^ and that diffusion-weighted MRI serves as an excellent, but time-limited, indicator of hypoxic-ischemic brain injury^7^, we hypothesized that apparent diffusion coefficient (ADC) values in the striatum and thalamus in the first few days of life could be quantitatively used to predict dystonia in CP following neonatal HIE.

## Methods

This retrospective case-control study was approved by the Washington University School of Medicine Institutional Review Board.

We examined records of all neonates born between 1/1/2010 and 12/31/2015 who underwent whole body therapeutic hypothermia for HIE (Table 1) at St. Louis Children’s Hospital, St. Louis, MO, USA (N = 243). MRIs were clinically-acquired at different time points for different subjects: 9 subjects had MRIs done on day 2, 2 subjects on day 3, 63 on day 4, and 69 on day 5. In patients undergoing therapeutic hypothermia, ADC values reach a nadir at approximately 4 days after injury and thereafter return to normal (pseudonormalize) by 12 days^7^. To optimize subject numbers while ensuring comparability between scans, we further examined only the 132 subjects with MRIs done on days 4–5 of life.

**Table 1.** Clinical inclusion and exclusion criteria for therapeutic hypothermia for neonatal hypoxic-ischemic encephalopathy (72 hours to core temperature of 33.5°C)

|  |  |
| --- | --- |
| Inclusion Criteria | ≥35 weeks' gestation<br>≤6 hours of age<br>Clinical encephalopathy (abnormal tone, poor responsiveness, or excessive irritability)<br>Known or suspected perinatal ischemic events evidenced by either catastrophic sentinel event or 2 criteria from the following: fetal or neonatal acidemia (pH <7.0 or based deficit >12 mEq/L), Apgar score of <5 at 5 minutes, or ventilatory support for ≥5 minutes. |
| Exclusion Criteria | Weight < 2 kg<br>Presence of major congenital abnormalities<br>Imminent Death |

Clinical MRI scans were performed on 3T Sonata and Avanto Siemens scanners (Erlangen, Germany). Diffusion sequences were obtained using a single-shot, echo planar imaging sequence with diffusion sensitization obtained in 3–12 different directions and b amplitudes of 0 and 1,000 s/mm^2^. Spatial resolution was 1.5 × 1.5 × 3 mm^3^. ADC values were determined using clinically-integrated software (IBM iConnect Access, Armonk, NY) in striatal and thalamic regions of interest (ROIs) drawn free-hand on single axial slices of ADC maps approximating Montreal Neurologic Institute (MNI) coordinate Z = 0. This simple and straightforward approach to generating ROIs lends itself to clinical application, but likely resulted in partial inclusion of surrounding structures. Also of note, the striatal ROI included the caudate and putamen traced together such that any intervening portion of the anterior limb of the internal capsule would also be included (Figure 1A). The lower of the two average ADC values from the left and right ROIs for each structure was used for further analysis (Figure 1A). ADC values were obtained by two investigators (K.C. and B.R.A), both blinded to motor phenotype.

**Figure 1.**
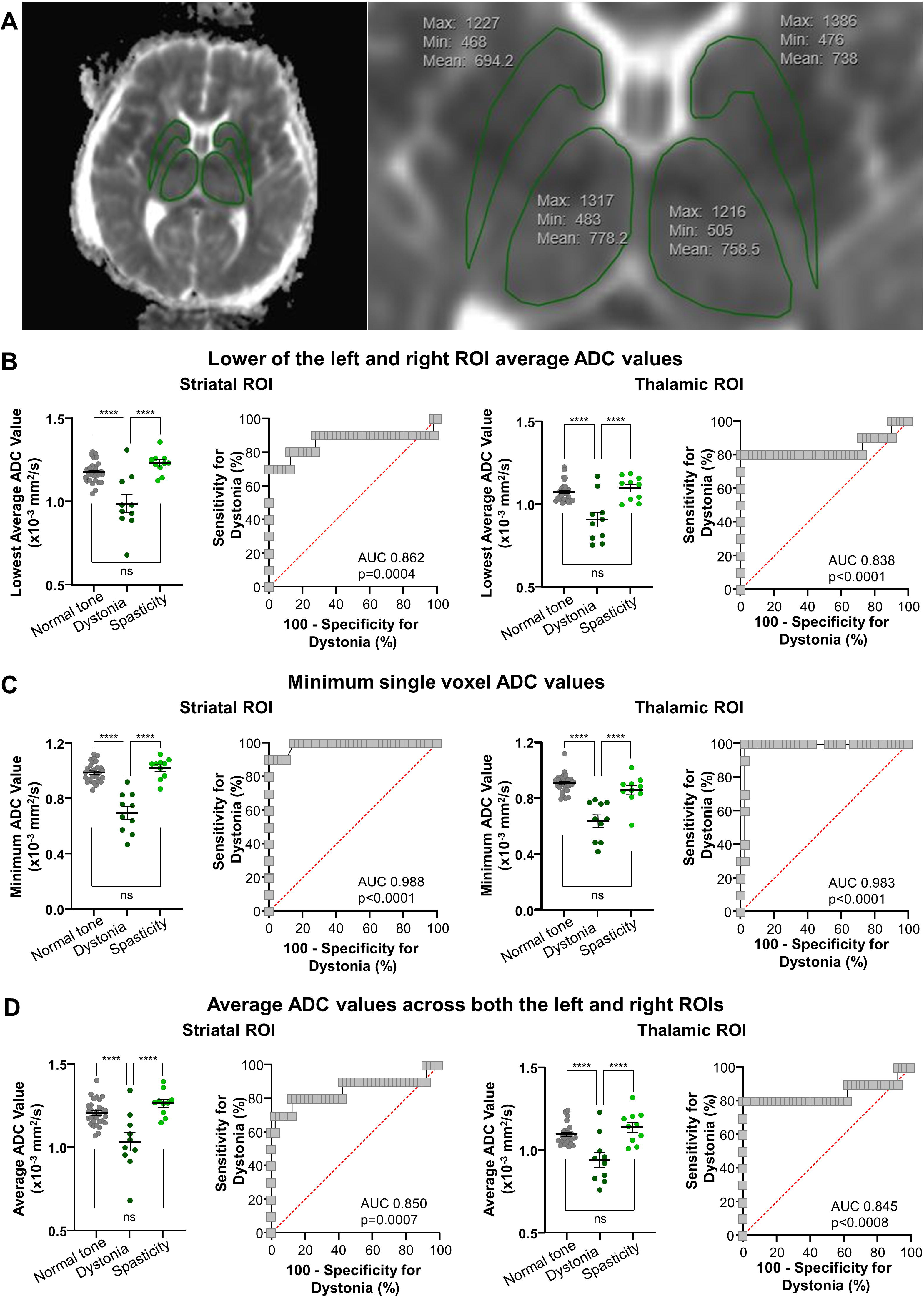
Striatal and thalamic apparent diffusion coefficient (ADC) values can be used to predict motor phenotypes. A) Example striatal and thalamic ROIs and ADC values in a subject who went on to develop dystonia. Based on the indicated ADC values, the lowest average ADC value between the left and right ROIs is 0.694 × 10^−3^ mm^2^/s for the striatum and 0.758 × 10^−3^ mm^2^/s for the thalamus. The single voxel minimum ADC value is 0.468 × 10^−3^ mm^2^/s for the striatum and 0.483 × 10^−3^ mm^2^/s for the thalamus. B) Comparison of striatal and thalamic lowest average ADC values between motor phenotypes. ADC values were significantly lower in subjects who go on to develop dystonia compared to those who develop spasticity without dystonia or have normal tone (****p 0.0001, one-way ANOVA with post-hoc Tukey HSD). Receiver operator characteristic curves demonstrate that striatal and thalamic ADC values were significant predictors of dystonia compared to spasticity alone or normal tone (AUC-area under the curve). Striatal ADC values less than 1.014 × 10^−3^ mm^2^/s were 100% specific and 70% sensitive for predicting dystonia. Thalamic ADC values less than 0.973 × 10^−3^ mm^2^/s were 100% specific and 80% sensitive for predicting dystonia. C) Comparison of striatal and thalamic single voxel minimum ADC values between motor phenotypes. Significant relationships were the same as when using lowest average ADC values, but with even higher ROC AUCs and improved sensitivity for predicting dystonia. Striatal minimum ADC values less than 0.846 × 10^−3^ s/mm^2^ were 100% specific and 90% sensitive for predicting dystonia. Thalamic minimum ADC values less than 0.780 × 10^−3^ s/mm^2^ were 98% specific and 90% sensitive for predicting dystonia. D) Comparison of striatal and thalamic single voxel average ADC values between motor phenotypes. Significant relationships were the same as when using lowest average ADC values, with comparable ROC AUCs but reduced sensitivity for predicting dystonia. Striatal minimum ADC values less than 1.060 × 10^−3^ s/mm^2^ were 100% specific and 60% sensitive for predicting dystonia. Thalamic minimum ADC values less than 0.976 × 10^−3^ s/mm^2^ were 100% specific and 70% sensitive for predicting dystonia.

Motor phenotypes were determined by chart review of neonatal neurologist, pediatric movement disorder specialist, and pediatric cerebral palsy specialist notes through 5 years old. Subjects were deemed to have CP and dystonia if they had persistent motor impairment^8^ and either explicitly documented dystonia or were described as having involuntary sustained postures that emerged or worsened with voluntary movement or heightened arousal.^9^ Of note, many of these subjects with dystonia also had some degree of spasticity, as it is well know that dystonia and spasticity commonly co-occur in CP.^10,11^ Subjects were deemed to have CP and spasticity (without dystonia) if this was explicitly documented or they were described as having hypertonia and hyperreflexia without any other evidence of dystonia^12^. Subjects with normal tone had serial notes documenting normal motor exams.

One-way ANOVA was used to compare ADC values between motor phenotypes. Receiver operator characteristic (ROC) curves were used to determine the sensitivity and specificity of ADC values for motor phenotype prediction (GraphPad Prism 8, San Diego, CA). Multinomial logistic regression was used to determine whether ADC values could predict motor phenotype when adjusting for key clinical variables (Table 1, SPSS, IBM, Armonk, NY).

## Results

Of the 132 subjects with MRIs obtained between days 4–5 of life, 82 subjects were excluded from further analysis because they were either lost to follow-up (33), did not have diffusion sequences acquired (1), had only documentation of hypotonia (21), or had only documentation of hypertonia without hyperreflexia or other clarifying features to allow for differentiation between spasticity and dystonia (27). Table 2 summarizes the clinical details for the included subjects (10 with spasticity without dystonia, 10 with dystonia, and 30 with normal tone). There was no missing clinical data for the included subjects.

**Table 2.** Subject details

| Motor Phenotype | Striatal Lowest Average ADC (x 10<sup>-3</sup> mm<sup>2</sup>/s) | Thalamic Lowest Average ADC (x 10<sup>-3</sup> mm<sup>2</sup>/s) | Striatal Minimum ADC (x 10<sup>-3</sup> mm<sup>2</sup>/s) | Thalamic Minimum ADC (x 10<sup>-3</sup> mm<sup>2</sup>/s) | Striatal Average ADC (x 10<sup>-3</sup> mm<sup>2</sup>/s) | Thalamic Average ADC (x 10<sup>-3</sup> mm<sup>2</sup>/s) | GA (wks) | Sex (% Male) | Birth Weight (g) | 5 min APGAR | Cord gas pH | Cord gas base deficit |
| --- | --- | --- | --- | --- | --- | --- | --- | --- | --- | --- | --- | --- |
| Normal tone (n=30) | 1.230 (1.190, 1.270) | 1.098 (1.053, 1.143) | 0.989 (0.965, 1.012) | 0.720 (0.688, 0.751) | 1.264 (1.217, 1.311) | 1.139 (1.079, 1.199) | 38.8 (38.2, 39.3) | 56.7 (47.6, 65.7) | 3370 (3160, 3570) | 4.93 (4.19, 5.68) | 7.09 (7.03, 7.14) | -10.0 (-13.1, -6.97) |
| Dystonia (n=10) | 0.989 (0.881, 1.096) | 0.907 (0.820, 0.995) | 0.698 (0.609, 0.787) | 0.520 (0.440, 0.599) | 1.033 (0.923, 1.142) | 0.941 (0.851, 1.030) | 39.0 (38.0, 40.0) | 50.0 (34.2-65.8) | 3330 (3107, 3555) | 3.10 (1.69, 4.52) | 7.05 (6.93, 7.16) | -15.5 (-20.1, -11.0) |
| Spasticity (n=10) | 1.177 (1.154, 1.199) | 1.074 (1.053, 1.095) | 1.021 (0.974, 1.067) | 0.684 (0.629, 0.739) | 1.204 (1.177, 1.231) | 1.094 (1.071, 1.117) | 37.6 (36.5, 38.7) | 70.0 (55.5, 84.5) | 3440 (3080, 3800) | 4.40 (3.09, 5.71) | 7.04 (6.92, 7.17) | -11.4 (-16.5, -6.22) |

Average values with 95% confidence interval lower and upper limits are indicated for each motor phenotype. ROI - region of interest, ADC-apparent diffusion coefficient, GA-gestational age, wks-weeks

When examining the lower of the two average ADC values from the left and right ROIs, subjects who developed dystonia had significantly lower ADC values in the striatum and thalamus when compared to subjects who developed spasticity or had normal motor development (p< 0.0001, one-way ANOVAs with post-hoc Tukey HSD, Table 2, Figure 1B). Striatal ADC values less than 1.014 × 10^−3^ mm^2^/s were 100% specific (95% CI 91–100%) and 70% sensitive (95% CI 40–89%) for predicting dystonia as opposed to spasticity or normal motor development with an ROC area under the curve (AUC) of 0.862 (95% CI 0.679–1.000, p = 0.0004). Thalamic ADC values less than 0.973 × 10^−3^ mm^2^/s were 100% specific (95% CI 91–100%) and 80% sensitive (95% CI 49–96%) for predicting dystonia with an AUC of 0.838 (95% CI 0.633–1.000, p = 0.001) (Figure 1B). Both striatal and thalamic ADC values remained significant predictors of motor phenotype even when adjusted for age at scan and scanner type (multinomial logistic regression, p< 0.0005).

A multinomial logistic regression model including ADC values in the striatum, ADC values in the thalamus, gestational age, sex, birth weight, 5 minute APGAR, cord gas pH, and cord gas base deficit significantly improved motor phenotype prediction accuracy from the null model (p = 0.004) with good model fit (Deviance p = 0.981). When adjusted for the other variables (including thalamic ADC value), only striatal ADC value remained a significant predictor of motor phenotype (p = 0.025).

Most images demonstrated diffusion restriction in small regions of each structure. To assess whether small regions of diffusion restriction contributed to the prediction of dystonia, we repeated the above analyses using the minimum single voxel ADC values for each structure. With the caveat that these single voxel values are susceptible to local artifact, using these values maintained all significant relationships obtained by using the lowest average ADC values but with even higher ROC AUCs and improved sensitivity for predicting dystonic CP (Figure

1C). We additionally examined the ADC value for each structure averaged across both the left and right ROIs (instead of taking the lower of the two average values from the left and right ROIs). Again, this measure also maintained all significant relationships described above with comparable ROC AUCs but lower sensitivity for predicting dystonic CP (Figure 1D). In sum, regardless of the method used to assess striatum and thalamic ADC values, dystonia could be predicted with high specificity.

## Discussion

ADC values in the striatum, perhaps more so than in the thalamus, obtained from MRIs done in 4–5 day old neonates show promise for predicting dystonia with high specificity following therapeutic hypothermia for HIE.

Predicting dystonia in CP has high clinical utility since dystonia requires different treatment from spasticity, is often under-recognized, and can be functionally debilitating^10^. Though deep gray matter injury has been subjectively associated with dystonia in CP^5^, this study is unique in that it demonstrates an objective and clinically-feasible method to assess the degree of deep gray matter injury that is most likely to result in dystonia. As the methods used here require only clinically-available image viewing software, basic neuroanatomical knowledge, and identification of the striatum and thalamus on only one axial slice on ADC maps, this approach can be easily integrated into clinical practice. Lending credence to the method we have described here, recently, a more generalized approach involving whole brain averaging of ADC values has been used for prediction of outcome in adult survivors of cardiac arrest.^13^ Our approach is additionally valuable due to its focus on motor phenotyping and the regional specificity of ADC measurements.

This study is limited by a number of factors. 1) It is retrospective. 2) A significant proportion of subjects were excluded due to lack of follow up or insufficient follow-up documentation of their motor phenotype, even though only notes of putative experts in motor phenotyping were included in our analysis. This speaks the to the clinical difficulty of differentiating between different forms of tone in CP,^10,14^ particularly in young children in whom valuable diagnostic aids like the Hypertonia Assessment Tool are not yet validated.^15–17^ 3) No single motor phenotyping expert examined all subjects. 4) Babies were imaged on two different MRI scanners. Though ADC values are theoretically independent of the MRI system, the success of this approach across scanners is encouraging regarding its generalizability.

Future prospective studies using ADC values to predict dystonic CP are needed in which designated experts examine each of the subjects for dystonia and spasticity in a standardized manner. It may be of additional value for these experts to gauge the predominant form of tone in these subjects as we could only gather retrospective documentation of the presence or absence of dystonia and not necessarily whether it is the primary issue limiting function in the subjects in our study. Therefore, rigorous documentation of functional ability using validated classification systems will be required to systematically characterize how these different tone abnormalities practically impact children.^18–22^ Noting that dystonia can be particularly functionally debilitating^10,23^, comparison of ADC values while accounting for functional status will be required to appropriately parse out whether the results described here predict the type of tone, the degree of functional impairment, or both. To this end, it is important to note that basal ganglia and thalamic lesions have been associated with functional impairment following neonatal brain injury although, as has the majority of literature in this area, this association is based on qualitative MRI review by expert consensus on not by using the quantitative and more clinically-feasible method demonstrated here.^1,2,6^ Still, despite its limitations, these results suggest that the practical and objective MRI assessment outlined here may serve to identify those babies for whom vigilance for dystonia following neonatal HIE is most indicated.

## Data Availability

We will make all data available to any qualified investigator upon request.

## What this paper adds

A clinically-feasible objective MRI method for dystonia prediction following neonatal hypoxicischemic encephalopathy motor abnormality documentation.

## Author Contributions

Ms. Chintalapati was responsible for design of the study, data acquisition and analysis, and data interpretation. Mr. Miao was responsible for data acquisition, analysis, and interpretation. Dr. Aravamuthan was responsible for the design and conceptualization of the study, data interpretation, and preparing the original draft of the manuscript. Dr. Mathur was responsible for data interpretation and revising the manuscript for intellectual content. Dr. Neil was responsible for design of the study, data interpretation and revising the manuscript for intellectual content.

## Study Funding

Funding supporting this work is from the National Institutes of Neurological Disorders and

Stroke (5K12NS098482–02).

